# Reservoir Computing in Epidemiological Forecasting: Predicting Chicken Pox incidence

**DOI:** 10.1101/2023.04.24.23289018

**Authors:** Kaushal Kumar

**Affiliations:** Institute for Applied Mathematics, Heidelberg University, Im Neuenheimer Feld, Heidelberg, 69120, BW, Germany

**Keywords:** Health Informatics, Chickenpox cases, Time Series Analysis, Deep Learning, Reservoir Computing

## Abstract

We examine the applicability of time-series forecasting techniques to model and predict chickenpox incidence rates using publicly available epidemiological data from Rozemberczki et al. [1]. Analyzing data across both time and location is crucial in understanding disease dynamics, allowing for the identification of patterns such as temporal clustering, detection of high-incidence areas, characterization of disease spread, measurement of temporal synchrony, and forecasting future incidence rates. The primary objective of this study is to evaluate the effectiveness of neural networks in addressing this problem. Reservoir Computing, ARIMA, and various types of Recurrent Neural Networks (RNNs) have demonstrated success in tackling complex time-series issues. We assess several models based on different RNN architectures, including Long Short-Term Memory (LSTM), Bidirectional LSTM (BLSTM), Gated Recurrent Unit (GRU), Bidirectional GRU (BGRU), and compare their performance. We use a deep learning approach based on Reservoir Computing to predict chickenpox counts based on past incidence rates. We implement all the aforementioned neural network architectures for forecasting chickenpox incidence rates and compare their prediction accuracy. Our results indicate that Reservoir Computing prediction models outperform all other models trained on the same data. Furthermore, we demonstrate that Reservoir Computing prediction models are more efficient and quicker to train and deploy in epidemiology.

## 1 Introduction

Preventing and controlling infectious diseases in a population is a critical task for public health organizations, and accurate quantitative prediction is essential for effective prevention and control. Epidemiological data is often recorded and distributed in bulk, which makes predicting future incidence based on previous measurements necessary. Health informatics data analysis plays a critical role in understanding disease dynamics by enabling the identification of space-time clustering, hot-spot detection, characterization of invasion waves, and quantification of spatial synchrony. In particular, spatial synchrony is significant for acute immunizing infections, as it can either permit or prevent regional persistence of infections, exacerbating the economic and public health burden, as seen with the COVID-19 outbreak.Here, our objective is to apply time-series forecasting techniques to model and predict the chickenpox case counts. To accomplish this goal, we analyze weekly case count data of chickenpox from all counties in Hungary and present the time-series plots of the data in Figure 1.

**Fig. 1.**
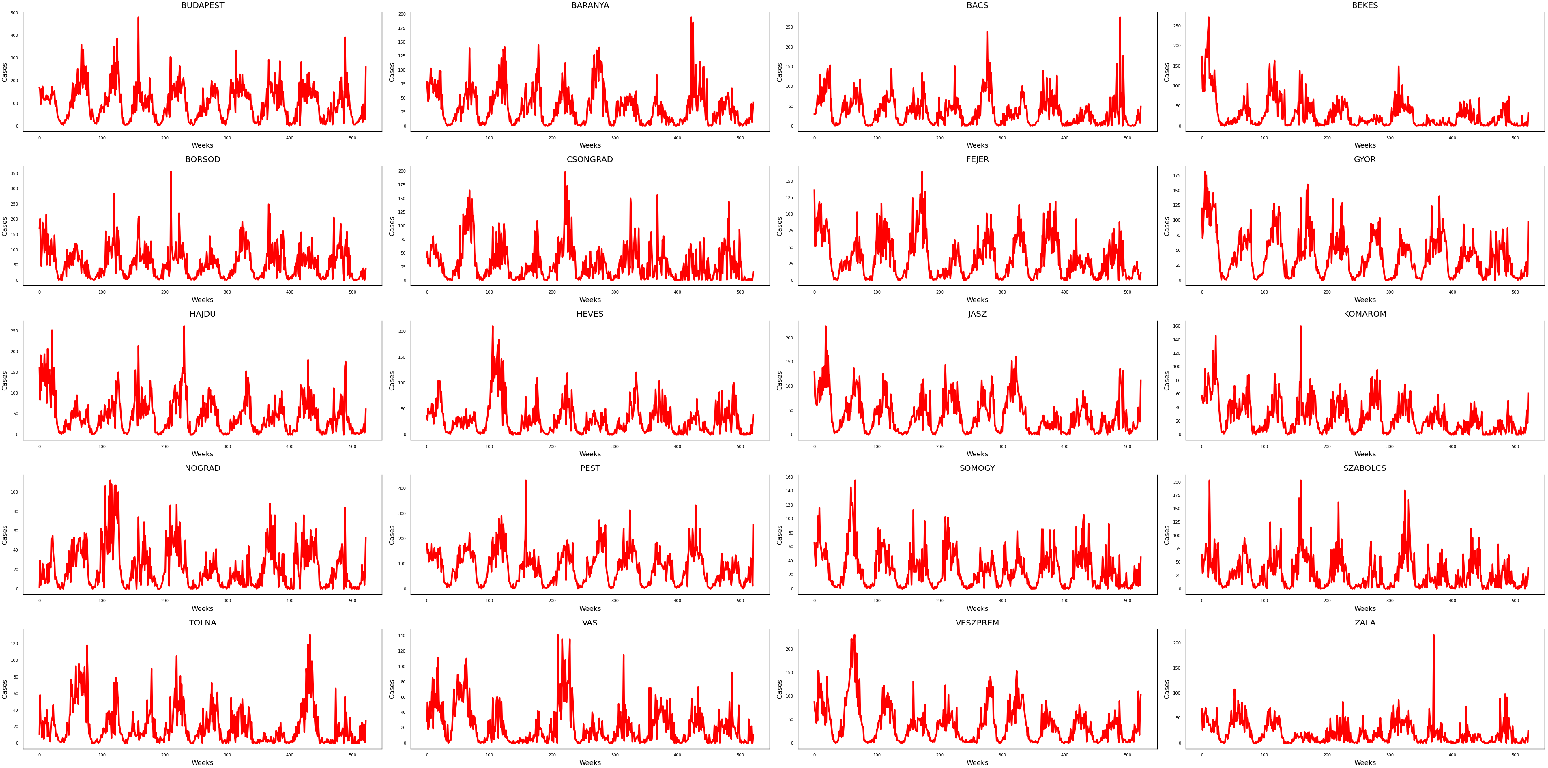
The above plots shows the time series of weekly chickenpox cases in a different location in Hungary. The dataset comprises a collection of time series representing the reported cases at the county level, spanning the period from 2005 to 2015.

Chickenpox is a highly contagious disease caused by the varicella-zoster virus (VZV), which poses a significant risk to vulnerable groups such as pregnant mothers, children, and individuals with compromised immune systems. According to the source [2], the majority of individuals in the European Union/European Economic Area (EU/EEA) typically acquire antibodies to VZV before the age of 10. However, there are variations in seroprevalence among countries, with children in southern and eastern Europe exhibiting lower seroprevalence rates compared to their counterparts in northern and western Europe. Such variations may be due to differences in the use of daycare and preschool facilities and social contacts. Newborns usually have maternal antibodies, which offer temporary protection against the virus.

Forecasting is a crucial task in many industries, and several methods have been developed to predict future outcomes [3] [4]. Commonly used methods include ARIMA, LSTM, GRU, Bidirectional LSTM, Bidirectional GRU, and Reservoir Computing. However, each method has its limitations. For example, ARIMA models perform well on small datasets and short-term predictions but struggle with large datasets and long-term predictions. Furthermore, computational efficiency is an important consideration, and some models may be more efficient than others.

Forecasting the spread of diseases is particularly challenging and requires specialized techniques. Deep learning techniques such as RNN, GRU, Reservoir Computing, and LSTM have shown promise in modeling sequential data and can be used for disease spread forecasting. Reservoir Computing models are better suited for forecasting complex systems such as nonlinear systems [5].

Choosing the right forecasting method depends on the specific task, dataset size, and desired prediction horizon. ARIMA models can be effective for small datasets and short-term predictions, while Reservoir Computing models are better for complex non-linear systems. Deep learning techniques such as RNN, GRU, Reservoir Computing, and LSTM are powerful tools for modeling sequential data and can be used for disease spread forecasting. Computational cost is also an important consideration, and the efficiency of the chosen model should be evaluated alongside its accuracy.

In this paper, we propose a three-level methodology for forecasting using deep learning models. The first step involves data pre-processing to normalize the data and ensure it falls within the range of [0, 1], making it suitable for network architectures and improving model accuracy. In the second step, the pre-processed data is split into training and testing sets, and deep learning models such as LSTM, GRU, and Reservoir Computing approach are trained on the training set using various hyperparameters. Finally, predictions are made on the test data using the trained models, and error matrices are analyzed to evaluate the performance of the models.

Overall, the proposed methodology provides a comprehensive approach for forecasting using deep learning models. By normalizing the data and using various deep learning architectures, the proposed methodology achieves accurate predictions. The results demonstrate the effectiveness of the proposed methodology for forecasting, and it has the potential to be applied to various fields, including finance, healthcare, and weather forecasting.

## 2 Related Work & Background

Time series analysis is an increasingly important research area that involves identifying patterns and trends in data that vary over time. There are various techniques available to analyze time series data, such as spectral analysis, wavelets, and autoregressive integrated moving average (ARIMA) models. Machine learning algorithms, including recurrent neural networks (RNNs) and long short-term memory (LSTM) networks, have also been applied to time series data analysis. The selection of a method depends on the data type and the specific research question.

Time series data is widely used in many fields, such as ecology, epidemiology, and physics, and is a collection of observations taken over a period of time that exhibit dependence. To analyze time series data, specialized statistics are employed, and it is represented in specific data structures that simplify further analysis.

Autocovariance and autocorrelation function (ACF) are essential measures of linear predictability and serial dependence in time series data. The autocovariance measures the covariance between two elements in the series, while the ACF is the Pearson correlation coefficient between two elements of a time series. Cross-correlation function (CCF) is another measure of linear predictability of one series from another. These measures help to identify patterns and relationships in time series data. In Figure 2 we determine the correlation between county case counts. As may be expected for a relatively small country with a good transportation network, there is a high degree of positive correlation between all counties, indicative of the ease of transmission of the disease.

**Fig. 2.**
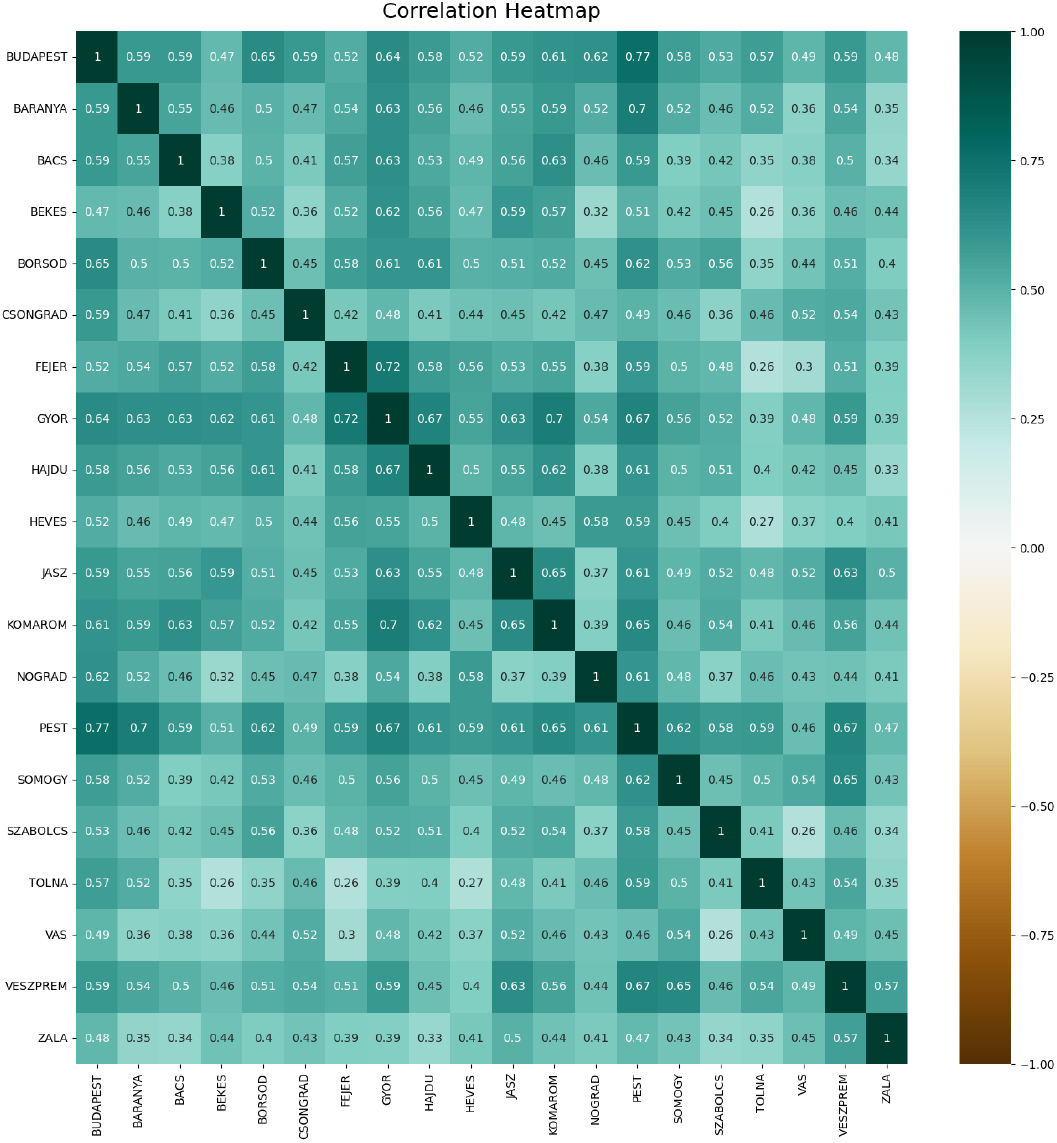
The above Heat-map shows the correlation matrix of the time series of weekly chickenpox cases in a different location in Hungary. There is a positive correlation between each other. One reason behind the positive correlation is that Hungary is a small country, and has good connectivity among all the counties, so the number of incidences increases.

Time series forecasting is crucial in various fields, including healthcare [6], finance, and meteorology. To address this problem, machine learning-based techniques have been proposed, such as ARIMA, LSTM, GRU, Bidirectional LSTM, Bidirectional GRU, and Reservoir Computing. These methods are useful in predicting future trends in time series data.

### 2.1 Autoregressive Integrated Moving Average

ARIMA stands for Autoregressive Integrated Moving Average [7]. It is a popular statistical method used for time series analysis and forecasting [8]. The ARIMA method essentially involves three steps:

- Autoregression (AR) - This step involves using past values of the time series to predict future values. In other words, the current value of the time series is assumed to be a linear function of its past values.
- Integrated (I) - This step involves differencing the time series to make it stationary, which means that its statistical properties do not vary over time.
- Moving Average (MA) - This step involves using past prediction errors to predict future values. In other words, the current value of the time series is assumed to be a linear function of its past prediction errors.

The ARIMA method can be used to model a wide range of time series data, including those with trends, seasonality, and irregular fluctuations. The method is particularly useful for short-term forecasting and can provide valuable insights into the underlying patterns and dynamics of the data [9]. However, it should be noted that the ARIMA method assumes that the data is stationary, and may not be suitable for all types of time series data. Additionally, it may not always provide accurate forecasts, especially for long-term predictions or when the underlying patterns in the data change over time [10].

### 2.2 Long Short-Term Memory

Long Short-Term Memory (LSTM) [11] [12] is a popular type of recurrent neural network (RNN) used for modeling sequential data. It addresses the problem of vanishing gradients in traditional RNNs by using memory cells that can store information over long periods of time. LSTMs have been applied successfully in various fields such as speech recognition, natural language processing, and time series prediction [13].

The key innovation of LSTM networks is the use of memory cells, which are units that can store information over time. These cells are connected to gates, which control the flow of information into and out of the cells. The gates are composed of sigmoid activation functions, which allow them to selectively filter the input data and decide what information to store and what information to discard [14].

LSTM networks are typically trained using backpropagation through time (BPTT), which is a variant of the backpropagation algorithm used in traditional feedforward neural networks. During training, the network is presented with a sequence of input data, and the weights of the network are updated to minimize the difference between the predicted outputs and the actual outputs [15].

The architecture of bidirectional LSTM is an extension of the traditional LSTM that allows information to be passed both forward and backward through the network. Bidirectional LSTMs have been shown to be effective in tasks such as speech recognition, language translation, and natural language processing.

In bidirectional LSTMs, the input sequence is processed in both forward and backward directions, and the hidden states of both directions are concatenated at each time step. This enables the network to capture information from both past and future contexts, which can improve prediction accuracy. However, the computational cost of bidirectional LSTMs is higher than traditional LSTMs because of the need to process the input sequence in both directions.

### 2.3 Gated Recurrent Unit

GRU is a type of recurrent neural network (RNN) that shares similarities with LSTM (Long Short-Term Memory) networks. Just like LSTM networks, GRU networks are designed to retain information over long periods of time, which makes them suitable for tasks such as speech recognition, language modeling, and time series prediction [16].

The key advantage of GRU networks is their computational efficiency as they use fewer gates compared to LSTM networks. GRU networks are designed with two gates - a reset gate and an update gate. The reset gate controls the amount of the previous hidden state that should be discarded, while the update gate determines the amount of new input to be added to the current hidden state [17].

During training, the weights of the network are updated using backpropagation through time (BPTT) to minimize the difference between the predicted outputs and the actual outputs. While GRU networks are suitable for applications where computational resources are limited, LSTM networks may outperform GRU networks for tasks that require modeling long-term dependencies in sequential data [18] [19].

In summary, GRU is a type of recurrent neural network that is similar to LSTM networks but with fewer gates, making it computationally more efficient. GRU networks have found applications in various fields, including natural language processing, speech recognition, and time series prediction.

Bidirectional GRU is a type of neural network architecture that is widely used for sequence modeling tasks such as natural language processing and speech recognition. Like a traditional GRU, it has two gates - a reset gate and an update gate. However, unlike a traditional GRU, a bidirectional GRU processes the input sequence in both forward and backward directions simultaneously.

This allows the model to capture past and future inputs, providing a richer representation of the input sequence. This can lead to improved performance on tasks where both past and future contexts are important. The output of a bidirectional GRU is the concatenation of the outputs from the forward and backward directions, which can be used for classification or regression tasks.

### 2.4 Reservoir Computing

Reservoir Computing [20][21](RC) is a relatively new approach to time series forecasting that is based on a recurrent neural network architecture [22]. The idea behind RC is to use the reservoir as a black box that maps input signals to output signals, without requiring explicit training of the recurrent connections in the network.

Two researchers first introduced the concept of RC in the early 2000s: Herbert Jaeger[23], who introduced echo-state networks (ESNs) using rate-coded neurons, and Wolfgang Maass[24], who introduced liquid state machines (LSMs) using spiking neurons.

Jaeger’s ESNs use a large pool of randomly connected neurons to create a reservoir that is driven by input signals. A linear output layer then reads out the reservoir dynamics to produce the network’s output. The reservoir is fixed during training, and only the output layer is trained to perform the desired task.

Maass’s LSMs use a similar approach, but with spiking neurons that communicate with each other through spikes. The reservoir is created by randomly connecting a large number of spiking neurons, and the input signals are fed into a subset of these neurons. The output of the network is then computed by measuring the activity of a readout neuron that receives inputs from a subset of the reservoir neurons. Again, the reservoir is kept fixed during training, and only the readout neuron is trained to perform the desired task.[25].

In summary, the above-mentioned time series forecasting methods offer different advantages and trade-offs. By implementing these methods on chickenpox case counts, the effectiveness and computational efficiency of each method can be evaluated, and the best method can be identified for this particular application.

## 3 Material Methods

This section provides an overview of the mathematical foundations underlying the time series techniques with Reservoir Computing (RC) [26] [27]. A basic architecture for RC is shown in Figure 3. This can be referred to as a temporal supervised learning task in the field of machine learning. It allows us the time series prediction that implements a desired mapping from input signals to output signals.

**Fig. 3.**
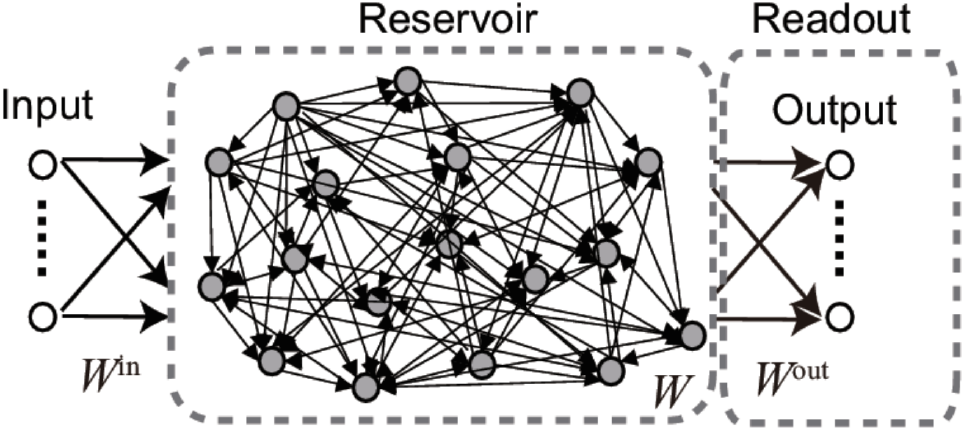
Structure of an echo-state network. Source [26].

### 3.1 Echo State Networks

Echo State Networks (ESNs) are a type of state-space models that utilize discrete-time, deterministic methods to predict a target output. Unlike traditional neural networks, ESNs have a trained output layer and stochastically generated input and hidden layers [23]. ESNs predict a target 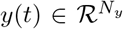 based on an input series 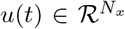, where *t ∈* [1, *T*], *T* is the length of the time series, *N*_*x*_ is the number of input series, and *N*_*y*_ is the number of output series being predicted. The hidden recurrent part of the ESN, known as the reservoir, consists of *N* neurons that act as a fading memory of previously processed inputs, summarized in the current state *x*(*t*).

At each time step *t* + 1, the ESN updates its state by receiving input *u*(*t*) and using the following equation:

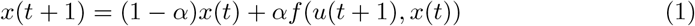

where *α* ∈ [0, 1] and *f* (*u, t*) = *tanh*(*W*_*in*_*u* + *W*.*x*) with *W*_*in*_ *∈* ℛ^*NS*^ as the input weights and *W ∈* ℛ ^*nN*^ as the weights of the hidden recurrent layer (the reservoir).

The ESN predicts the target at time *t* + 1 using:

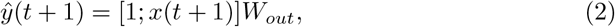

where *W*_*out*_ ∈ ℛ are the output (readout) weights of the network, and the concatenated column of ones acts as the bias.

ESN construction involves two stages. In the first stage, the input weights *W*_*in*_ and the hidden weights *W* are generated stochastically and remain fixed. In the second stage, the readout weights *W*_*out*_ are optimized. To obtain *W*_*out*_, all the states in *X* are collected, and the corresponding target *y* is given. The least squares problem is then minimized with a regularization parameter *λ* >= 0, which is expressed as:

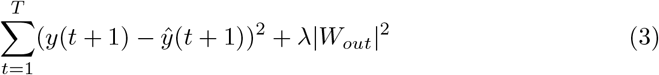

where λ >= 0 is a regularization parameter.

### 3.2 Forecasting accuracy measures

The Root-Mean-Square Error (RMSE) is commonly used to evaluate the accuracy of a model’s predictions. It assesses the variance or residuals between predicted and actual values and is utilized to compare the prediction errors of various models for a given dataset, rather than across datasets. The formula for calculating RMSE is as follows:

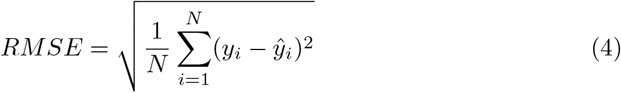

Where *N* represents the total number of observations, *y*_*i*_ represents the actual value, and ŷ _*i*_ represents the predicted value. One of the primary advantages of using RMSE is that it places greater weight on significant errors while scaling the scores in the same units as the forecast values (i.e., per week for this study).

## 4 Results

### 4.1 Data sets

The dataset [1] [28] comprises a collection of time series representing the reported cases at the county level, spanning the period from 2005 to 2015. The dataset is designed to facilitate two interrelated tasks, namely: forecasting of the case count at the county level and forecasting of the case count at the national level. The size of the dataset is (521, 20).

### 4.2 Data pre-processing

Data are normalized using Eq. (8). The range of the normalized data is [0, 1].

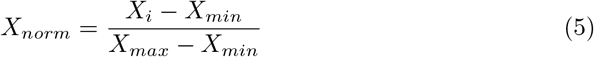

where *X*_*norm*_, *X*_*i*_, *X*_*min*_, and *X*_*max*_ are the normalized, observed, minimum, and maximum values of chickenpox cases per week.

### 4.3 Training and evaluation

Before initiating the training process, the dataset was divided into two subsets: a training set and a validation set. The training set was utilized to teach the system, while the validation set was used to evaluate the system’s performance. This technique was implemented to guarantee that the system could apply the acquired knowledge to new, unseen data. Assessing the performance of the system solely based on the training data would only measure memorization instead of learning. To maintain a consistent division, 80% of the data was allocated for training, and 20% was reserved for validation. It’s important to note that the splitting was done thoughtfully to ensure the reliability and validity of the results.

### 4.4 Analysis

In our setup, the Reservoir parameter is defined as the number of recurrent units = 20, leaking rate = 0.75, spectral radius (rho) = 0.9, and Scaling of input matrix= 1.0, we choose the connectivity, which defines the probability that two neurons in the reservoir are being connected (the two neurons can be a neuron and itself) Connectivity of recurrent matrix *W* = 0.15, Connectivity of input matrix = 0.2, Connectivity of feedback matrix = 1.1, and regularization coefficient = 1*e –* 8.

The outcomes of the study are effectively communicated through the use of Tables 1, 2, and 3, which are included in the results section of this paper. The analysis of chickenpox case count time series data using different models revealed that both the ARIMA model and the Reservoir Computing approach were effective in predicting the incidence of chickenpox. However, the Reservoir Computing approach demonstrated the lowest overall Rooted Mean Squared Error (RMSE) value compared to other models.

**Table 1.** RMSE for different Models

| Counties of Hungary | ARIMA | LSTM | GRU | BLSTM | BGRU | Reservoir Computing |
| --- | --- | --- | --- | --- | --- | --- |
| BUDAPEST | 0.1183 | 0.1852 | 0.1891 | 0.1846 | 0.1956 | 0.1145 |
| BARANYA | 0.1588 | 0.2153 | 0.2194 | 0.2160 | 0.2219 | 0.1544 |
| BACS | 0.1413 | 0.1616 | 0.1583 | 0.1682 | 0.1587 | 0.1346 |
| BEKES | 0.0607 | 0.0874 | 0.0884 | 0.0862 | 0.0899 | 0.0615 |
| BORSOD | 0.1018 | 0.1558 | 0.1567 | 0.1602 | 0.1656 | 0.0995 |
| CSONGRAD | 0.1322 | 0.1696 | 0.1745 | 0.1757 | 0.1740 | 0.1324 |
| FEJER | 0.1041 | 0.1390 | 0.1494 | 0.1496 | 0.1455 | 0.1041 |
| GYOR | 0.1159 | 0.1650 | 0.1620 | 0.1649 | 0.1733 | 0.1110 |
| HAJDU | 0.1294 | 0.1965 | 0.1945 | 0.2098 | 0.1940 | 0.1265 |
| HEVES | 0.0964 | 0.1390 | 0.1428 | 0.1417 | 0.1468 | 0.0987 |
| JASZ | 0.1097 | 0.1808 | 0.1759 | 0.1766 | 0.1704 | 0.1054 |
| KOMAROM | 0.0710 | 0.1036 | 0.1076 | 0.1031 | 0.1096 | 0.0768 |
| NOGRAD | 0.1264 | 0.2068 | 0.2204 | 0.2194 | 0.2101 | 0.1298 |
| PEST | 0.1043 | 0.1874 | 0.1806 | 0.1893 | 0.1849 | 0.1038 |
| SOMOGY | 0.1330 | 0.1866 | 0.1816 | 0.1805 | 0.1898 | 0.1360 |
| SZABOLCS | 0.1031 | 0.1415 | 0.1374 | 0.1356 | 0.1399 | 0.0993 |
| TOLNA | 0.1416 | 0.2676 | 0.2664 | 0.2718 | 0.2721 | 0.1612 |
| VAS | 0.1082 | 0.1489 | 0.1441 | 0.1504 | 0.1447 | 0.1079 |
| VESZPREM | 0.0890 | 0.1405 | 0.1315 | 0.1397 | 0.1401 | 0.0887 |
| ZALA | 0.0816 | 0.0971 | 0.0965 | 0.0968 | 0.0992 | 0.0811 |

Source: This is an example of table footnote. This is an example of table footnote.

**Table 2.** Run Time

| Methods | ARIMA | LSTM | GRU | BLSTM | BGRU | Reservoir Computing |
| --- | --- | --- | --- | --- | --- | --- |
| Time (seconds) | 6.75 | 83.02 | 85.06 | 60.29 | 55.90 | 0.67 |

The above Table shows the values of computational cost (time taken for run time) of weekly chickenpox cases in Budapest.

**Table 3.** RMSE and Run Time for Country Level

| Methods | ARIMA | LSTM | GRU | BLSTM | BGRU | Reservoir Computing |
| --- | --- | --- | --- | --- | --- | --- |
| RMSE | 0.1048 | 0.2119 | 0.2253 | 0.2089 | 0.2052 | 0.1041 |
| Time (seconds) | 7.68 | 80.78 | 79.20 | 59.92 | 57.90 | 0.74 |

The above Table shows the values of RMSE and computational cost of weekly chickenpox cases at the overall country level in Hungary.

Figure 4 showcases the prediction results for chickenpox case counts in Budapest, with the blue line representing the actual counts and the red line indicating the predicted cases. The study was extended to all counties in Hungary, and the RMSE values were calculated and presented in Table 1. Among all the models, the Reservoir Computing approach performed the best. Additionally, Table 2 shows that the Reservoir Computing approach required less time and was faster than the other models. In Figure 5, we predict the red curves to show the future case counts for the next 100 weeks in Budapest i.e, from week 521 to 620 is a future forecast of the chickenpox counts in Budapest.

**Fig. 4.**
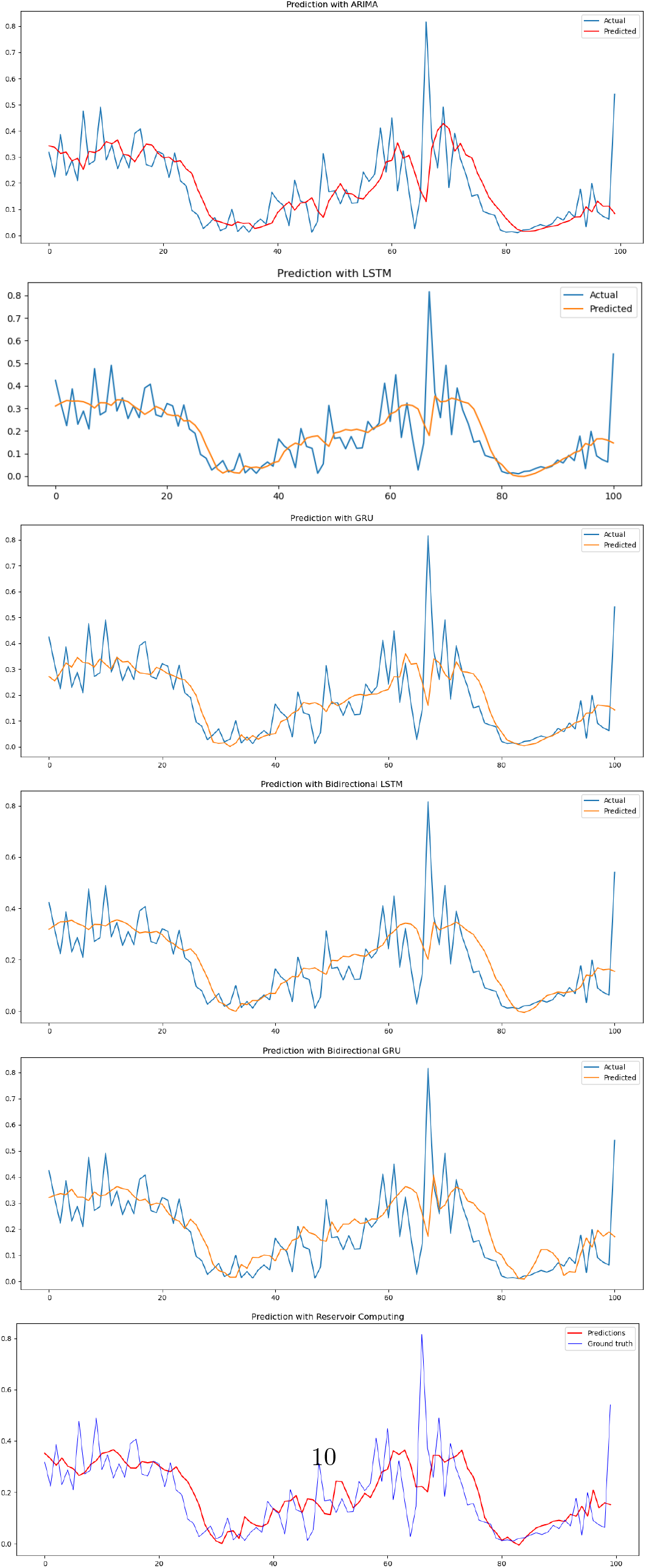
The above plots shows the time series prediction of weekly chickenpox cases in Budapest using a different approach.

**Fig. 5.**
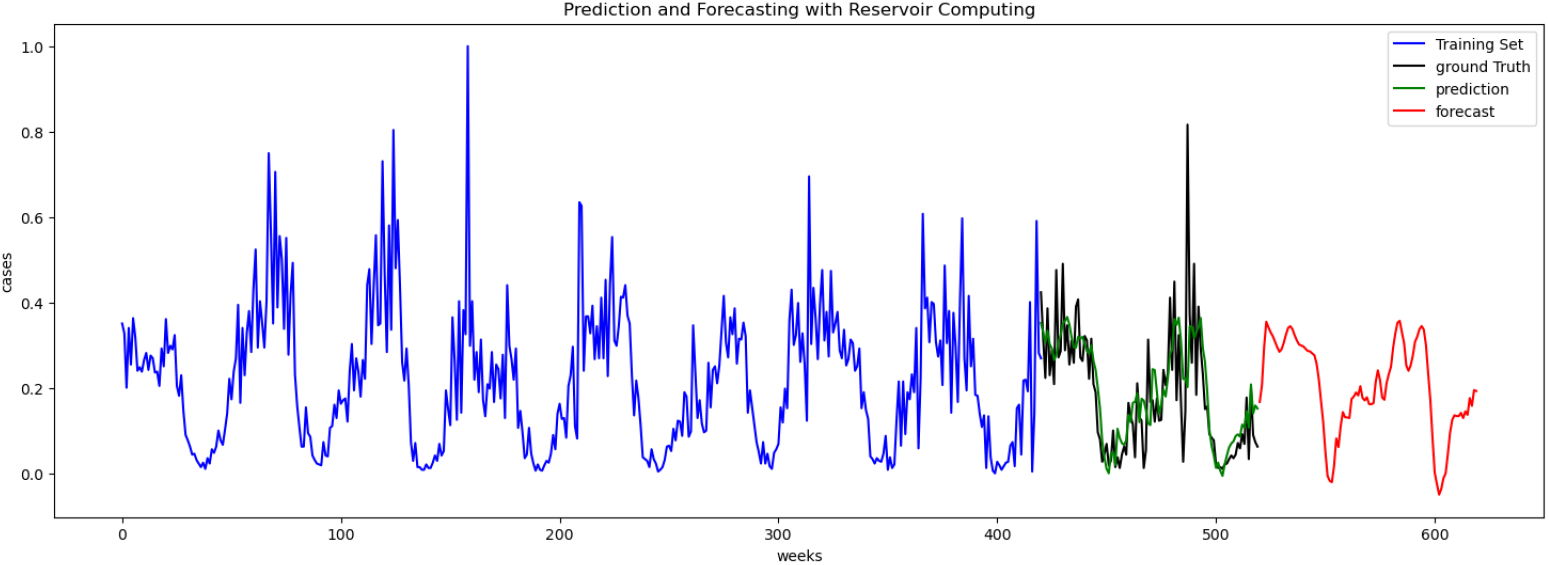
The above plots shows the time series prediction of weekly chickenpox cases in Budapest using the Reservoir Computing approach. The red curves show the future case counts for the next 100 weeks.

To predict the number of cases at the national level, the weekly cases from different regions were aggregated, and the same methodology was followed for the county-level data. Figure 6 depicts the actual versus predicted cases, while Table 3 shows the errors between the actual and predicted values. Consistent with earlier findings, the Reservoir Computing approach outperformed the other models in terms of accuracy and computational time. In Figure 7, we predict the red curves to show the future case counts for the next 100 weeks at the country level i.e, from week 521 to 620 are a future forecast of the chickenpox counts at the country levels.

**Fig. 6.**
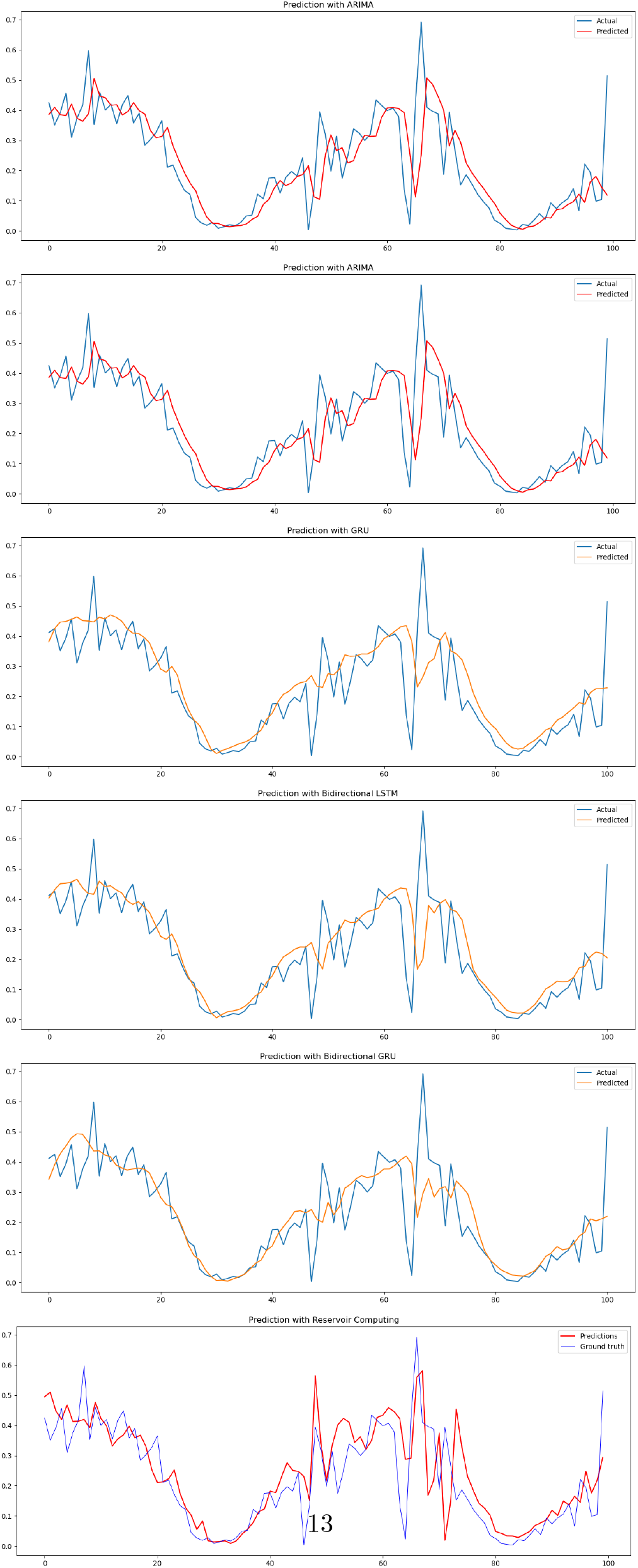
The above plots shows the time series prediction of weekly chickenpox cases at the overall country level in Hungary using a different approach.

**Fig. 7.**
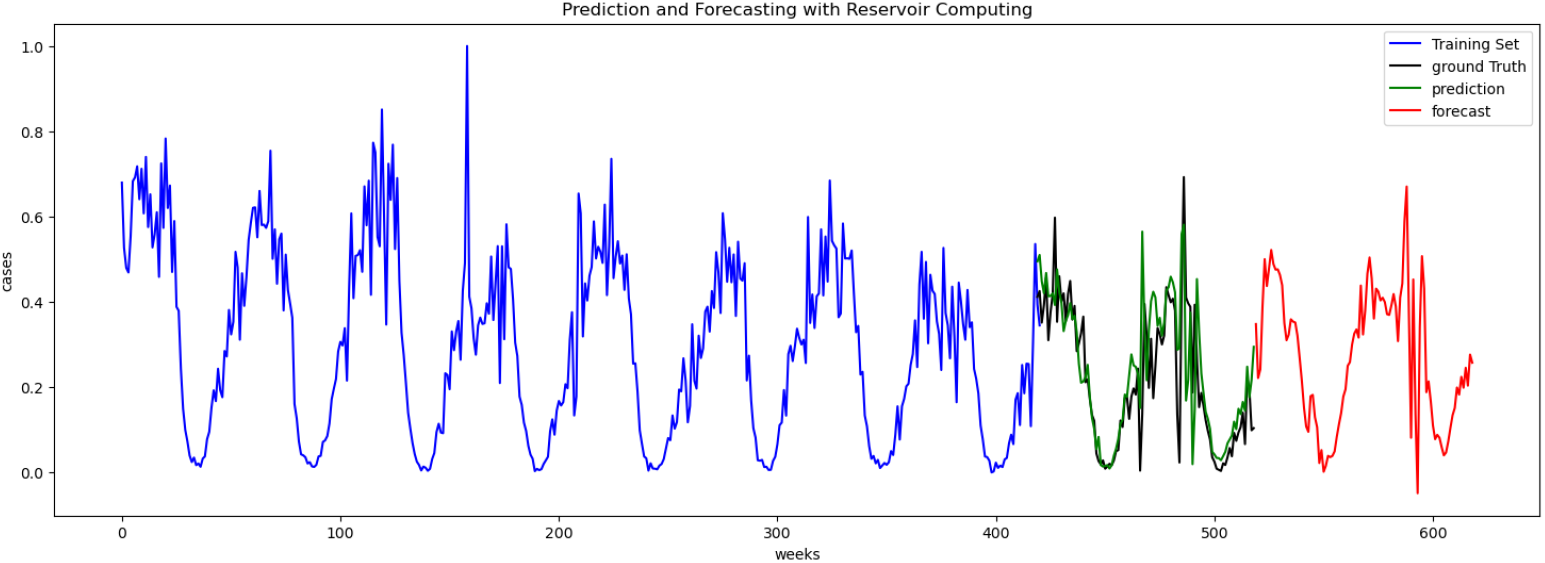
The above plots shows the time series prediction of weekly chickenpox cases at the overall country level in Hungary using the Reservoir Computing approach. The red curves show the future case counts for the next 100 weeks.

## 5 Conclusion

The increasing popularity of machine learning-based and deep learning algorithms in various fields has led to a growing interest in comparing their accuracy and effectiveness with traditional methods. In this study, we compared four representative techniques, namely ARIMA, LSTM, GRU, and Reservoir Computing, for forecasting time series data using chickenpox case count data. Our results demonstrate that the Reservoir Computing approach outperformed all other methods in terms of both prediction accuracy and computational efficiency. However the result suggests that ARIMA and RC are comparable, but ARIMA will be worse for larger data sets and take more time in computations. These interventions have the potential to offer valuable insights and support to governments, organizations, and individuals, enabling them to effectively minimize the spread and impact of epidemics, as well as prepare for and respond to any future incidence.

These results highlight the potential benefits of applying deep learning-based algorithms and techniques for predicting and forecasting health informatics and epidemiology-related data. We plan to extend our research to other prediction problems and datasets with varying numbers of features. Overall, our study contributes to the ongoing discussions around the effectiveness of machine learning-based and deep learning algorithms in various fields, particularly in health informatics and epidemiology.

## Data Availability

Public dataset

https://archive.ics.uci.edu/ml/datasets/Hungarian+Chickenpox+Cases

## Author contribution

K.K conception and design of the study, data collection, development of the machine learning models, analysis, and drafting of the article.

## Availability of data and materials

Data used in this study is publicly available [28].

## Code availability

The code associated with this study will be available upon acceptance of the manuscript on GitHub ^1^. For any additional information or clarification on the results presented, the corresponding author can be contacted via email upon reasonable request.

## Footnotes

1 https://github.com/kaushalkumarsimmons/Chickenpox-Time-Series-Analysis

